# Prevalence and patterns of ocular injury among patients admitted with head injury at a tertiary referral hospital in Uganda

**DOI:** 10.64898/2026.08.06.26359857

**Authors:** Alfred Olupot, Faith Oguttu, Joshua Shiuma, Oscar Jude Lyazzi, Bernard Odong, Mlaluko Rajabu Jumanne, Kamya Frank, David Mukunya, Ssali Grace Nsibirwa, Immaculate Atukunda

**Affiliations:** Department of Ophthalmology, Makerere University College of Health Sciences, Kampala, Uganda; Department of Community and Public Health, Busitema University, Mbale, Uganda

**Keywords:** traumatic brain injury, head trauma, ocular trauma

## Abstract

**Background:** Ocular injuries concomitantly occur among patients with head trauma because of the close anatomical relationship between the cranium and the orbit. Unfortunately, they are often overlooked during acute trauma care and may result in preventable visual loss. There is a dearth of data describing the burden and spectrum of ocular injury among head injury patients in sub-Saharan Africa. We conducted a hospital-based cross-sectional study to determine the prevalence and describe the patterns of ocular injury among adults admitted with head injury at the Accidents and Emergency Unit of Mulago National Referral Hospital, Uganda, from 15^th^ May 2025 to 30^th^ July 2025.

**Methods:** Consecutive patients aged 18 years and above with a confirmed head injury diagnosis underwent a structured ophthalmic evaluation comprising visual acuity testing, tonometry, slit-lamp and dilated fundus examination, pupillary and ocular motility assessment, confrontation visual fields, and review of craniofacial computed tomography. Ocular injuries were classified by anatomical site and injury type, and the prevalence was reported with a 95% confidence interval.

**Results:** Of 383 patients evaluated (mean age 32.7 ± 12.3 years; 89.3% male), road traffic accidents were the leading mechanism of injury (68.9%) and most patients had mild head injury (72.3%). Overall, 268 patients (70.0%; 95% CI 65.1-74.5) sustained at least one ocular injury. Adnexal injuries were most frequent (64.0%), dominated by periorbital oedema (45.2%), subconjunctival haemorrhage (42.3%) and eyelid ecchymosis (33.2%); orbital fractures occurred in 27.4%. Closed and open globe injuries were present in 13.8% and 0.8% of patients, respectively, and abnormal confrontation visual fields in 28.2%.

**Conclusion:** Ocular injury is highly prevalent among patients admitted with head injury in this setting, with adnexal and orbital structures most commonly affected. Routine ophthalmic assessment should be integrated into the initial evaluation of all head injury patients to reduce missed injuries and prevent avoidable visual morbidity.

## Introduction

Head injury is a major contributor to trauma-related morbidity and mortality worldwide and accounts for a substantial proportion of emergency department visits and hospital admissions [1]. The burden falls disproportionately on low- and middle-income countries (LMICs), where rapid urbanization, increasing motorization and limited enforcement of road safety have driven persistently high rates of road traffic injury [2, 3]. In Sub-Saharan Africa, head injury affects predominantly young, economically productive adults, with considerable personal, social and economic consequences [4, 5].

The eye and its adnexa are particularly vulnerable during craniofacial trauma owing to their exposed position and proximity to the facial skeleton and skull base [6]. Traumatic forces are transmitted from the skull through the orbital bones to the ocular structures, so that injuries may involve the adnexa, orbit, globe, optic nerve or visual pathways. They range from minor, self-limiting conditions such as subconjunctival haemorrhage to severe, sight-threatening injuries including globe rupture, posterior segment trauma and traumatic optic neuropathy [7, 8].

Early recognition of ocular injury in head injury patients is critical because delayed diagnosis may lead to permanent visual impairment or blindness [9]. In acute trauma settings, however, ocular injuries are frequently overlooked. Reduced consciousness, periorbital swelling, competing neurological injuries and limited ophthalmic expertise all contribute, and these challenges are amplified in resource-limited settings, where patients may be discharged or transferred without an adequate ocular assessment [9, 10].

Evidence from high-income countries describes the prevalence and spectrum of ocular injuries associated with head trauma and underscores the value of multidisciplinary care and early ophthalmology involvement [11, 12]. Data from Sub-Saharan Africa remain limited and are largely confined to small series or to studies restricted to specific injury mechanisms. Prior work at Mulago National Referral Hospital (MNRH) examined patients injured in road traffic accidents or reported visual deficits after traumatic brain injury, but did not comprehensively characterize the full anatomical spectrum of ocular injury across all head-injury mechanisms [13, 14].

Understanding the prevalence and distribution of ocular injuries in this population is essential for clinical practice and health policy. Such data support the integration of ophthalmic screening into trauma protocols, guide the training of non-ophthalmic clinicians and inform context-appropriate referral pathways. We therefore aimed to determine the prevalence and describe the patterns of ocular injury among patients admitted with head injury at MNRH, Uganda.

## Materials and methods

### Study design

We conducted a hospital-based, quantitative cross-sectional study among adults admitted with head injury.

### Study setting

The study was carried out from 15^th^ May 2025 to 30^th^ July 2025 at the Accidents and Emergency Unit of MNRH, Kampala, Uganda. Founded in 1913 and located on Mulago hill in Kampala, MNRH is Uganda’s national tertiary referral hospital and the teaching hospital of Makerere University College of Health Sciences. It has an established capacity of about 1,500 beds and admits an estimated 130,000-140,000 patients annually. The Accidents and Emergency Unit has a 59-bed capacity, comprising a resuscitation room, a general trauma section, radiology and laboratory services and three operating theatres, and is staffed by nurses, medical officers, neurosurgeons, orthopaedic and general surgeons, a trauma surgeon, an anaesthesiologist and residents. The unit receives a high volume of trauma cases from across the country, including most of the severe head injuries referred from lower-level facilities, and patients with head injury routinely undergo brain computed tomography (CT) within the hospital complex.

### Study population and eligibility

Consecutive adult patients (aged ≥ 18 years) admitted with a confirmed clinical diagnosis of head injury during the study period (May to July 2025) were eligible. Both conscious and unconscious patients were enrolled. Patients with ocular trauma but without a history of head injury were excluded. For conscious patients, written informed consent was obtained; for unconscious patients, consent was provided by the primary caregiver, and where the patient was unconscious but had no primary caregiver available, a waiver of initial consent was obtained from the institutional review board with deferred consent sought once the patient regained consciousness or a caregiver was traced.

### Sample size

The sample size was calculated using the Kish-Leslie formula for prevalence studies, assuming a prevalence of ocular injury of 34.6% derived from a previous study of road traffic accident victims at MNRH [13], a 95% confidence level and 5% precision. This yielded 348 participants; after adjusting for a 10% non-response rate, the final target sample size was 383. Sample sizes computed for the secondary objectives were smaller, so the prevalence estimate determined the final sample size.

### Data collection and ophthalmic assessment

Sociodemographic and clinical data were collected using a structured questionnaire administered through Kobo Toolbox (S1 File). After admission and stabilization, eligible patients or their caregivers were approached by trained research assistants for screening, consent and data collection. All participants underwent a systematic ophthalmic evaluation performed by the principal investigator, conducted, where the patient’s condition allowed, in the following sequence: inspection in ambient light; distance visual acuity using a 3-metre Snellen chart (or the illiterate “E” chart) recorded in Snellen notation; in patients who were unconscious and unable to cooperate with standard acuity testing, visual acuity was assessed indirectly using the pupillary light reflex, with a positive direct pupillary response to a bright light stimulus recorded as light perception and an absent response recorded as no light perception; intraocular pressure measurement using a Schiötz tonometer (average of three readings; categorized as low < 10 mmHg, normal 10-21 mmHg, high > 21 mmHg); portable slit-lamp examination of the anterior segment with fluorescein staining and the Seidel test; assessment of pupillary responses including the swinging-flashlight test for a relative afferent pupillary defect; evaluation of ocular motility, and alignment using the Hirschberg corneal-reflex test; and dilated fundus examination by indirect ophthalmoscopy (Keeler Vantage Plus) after instillation of tropicamide 0.5%. Topical proparacaine 0.5% was used to relieve pain and aid lid opening. Confrontation visual fields were assessed in conscious, cooperative patients. The orbital rim was palpated for fractures and the orbit for emphysema, and craniofacial CT images were reviewed for orbital, cranial and facial fractures, orbital emphysema, retrobulbar haemorrhage and rectus-muscle entrapment. The order of examination was adapted as necessary according to the patient’s level of consciousness and the severity of injury.

### Definition and classification of ocular injury

Ocular injury was defined as any traumatic abnormality involving the ocular adnexa, globe, orbit, optic nerve or the cranial nerves supplying the ocular system. Injuries were categorized by anatomical site (adnexal, anterior segment, posterior segment and neuro-ophthalmic) and by injury type. Globe injuries were classified according to the Birmingham Eye Trauma Terminology (BETT) system as closed globe (intact corneoscleral wall) or open globe (full-thickness corneoscleral defect) [15, 16].

### Statistical analysis

Data were exported from Kobo Toolbox, cleaned and analyzed using Stata version 18.0 (StataCorp, College Station, TX, USA). Frequencies, proportions and means with standard deviations were computed. Age was grouped into clinically relevant categories [17]. The prevalence of ocular injury was calculated as the proportion of enrolled patients with at least one ocular injury and reported with a 95% confidence interval (CI). Patterns of ocular injury were summarized by anatomical site and injury type as frequencies and percentages of the total cohort.

### Ethical considerations

Ethical approval was obtained from the School of Medicine Research and Ethics Committee (SOMREC) of Makerere University College of Health Sciences and the institutional review board of MNRH, and the study was registered with the Uganda National Council for Science and Technology (study number MAK-SOMREC-1254). The study adhered to the principles of the Declaration of Helsinki. Written informed consent was obtained from participants or their legally authorized representatives before enrolment; participants who could not write indicated consent with an inked right-thumb print. For unconscious patients without an available caregiver, a waiver of initial consent was granted by the institutional review board and deferred consent was obtained once the patient regained consciousness or a caregiver was traced. Confidentiality was maintained by identifying participants with a four-digit study number, and all patients found to have ocular injuries received appropriate ophthalmic management.

## Results

### Participant characteristics

During the study period, 1,784 patients were admitted to the Accident and Emergency Unit of MNRH. Of these, 1,026 were admitted for reasons other than head injury (e.g., fractures, hernias) and were excluded, leaving 758 patients admitted with a confirmed head injury. Among these, 56 were children, 6 adults declined consent, 5 adults died before enrolment, and 308 patients were admitted at night or over the weekend when the study team was not available; the remaining 383 adults met the inclusion criteria and were enrolled (Fig 1). The mean age was 32.7 ± 12.3 years (range 18-83) and most participants were male (89.3%). Business/ self-employment (27.4%) and commercial motorcycle riding (boda-boda) (25.1%) were the most frequent occupations, most participants had attained primary level education (53.5%) and resided in urban areas (65.5%). 5% were known to have used head-protective equipment at the time of injury.

**Fig 1.**
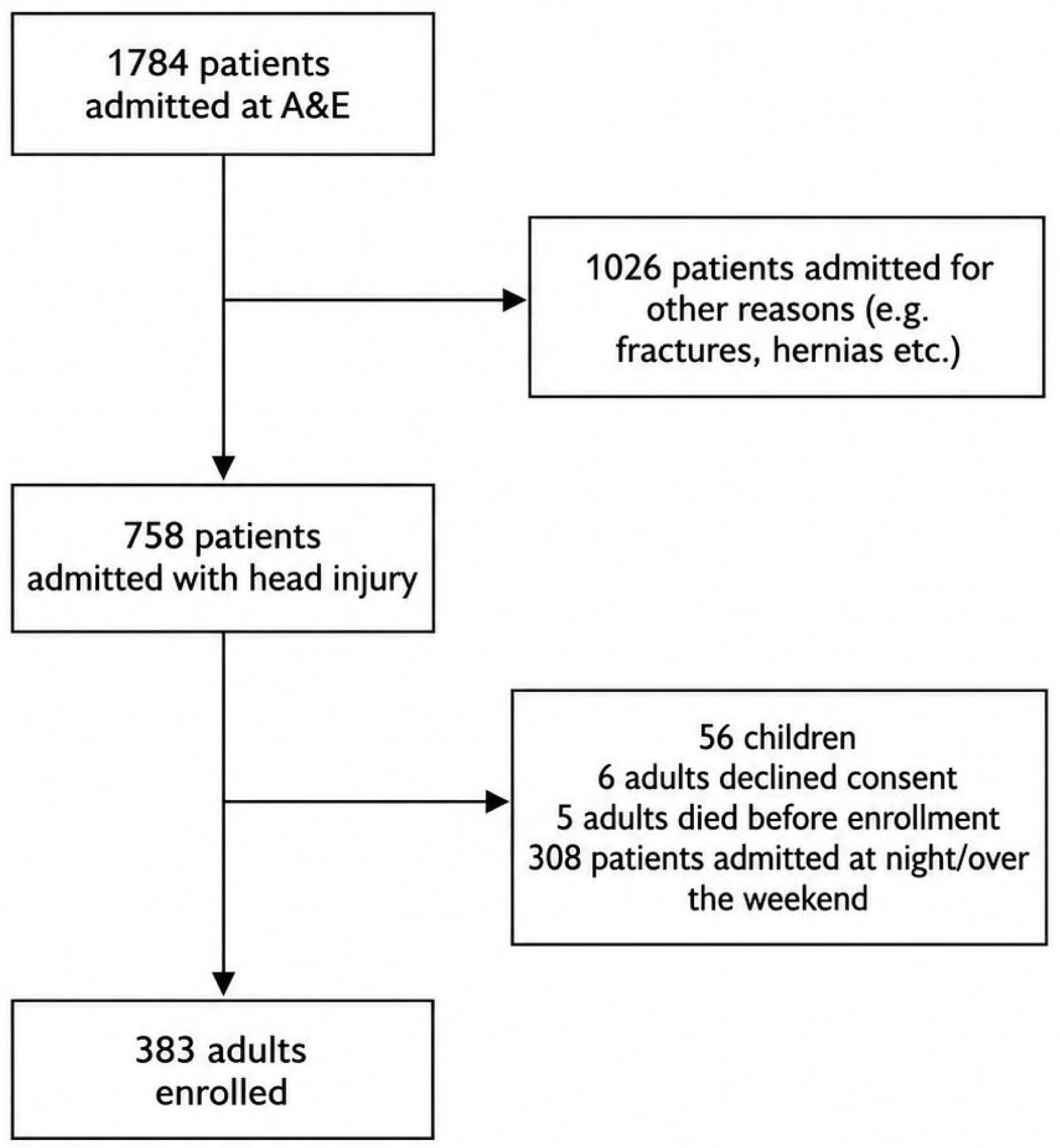
Flowchart of participant screening and enrolment at the Accidents and Emergency Unit of Mulago National Referral Hospital (MNRH), Uganda, May-July 2025. Of 1,784 patients admitted during the study period, 758 had a confirmed head injury; after excluding 56 children, 6 adults who declined consent, 5 adults who died before enrolment, and 308 patients admitted at night or over the weekend, 383 adults met the inclusion criteria and were enrolled.

Road traffic accidents were the predominant mechanism of injury (68.9%), followed by assault (23.5%), and most patients had mild head injury on the Glasgow Coma Scale (72.3%). Facial and cranial fractures were present in 46.7% and 53.3% of patients, respectively, and cerebral contusion was the most common intracranial haemorrhagic lesion (58.6%). Participant characteristics are summarized in Tables 1 and 2.

**Table 1:**
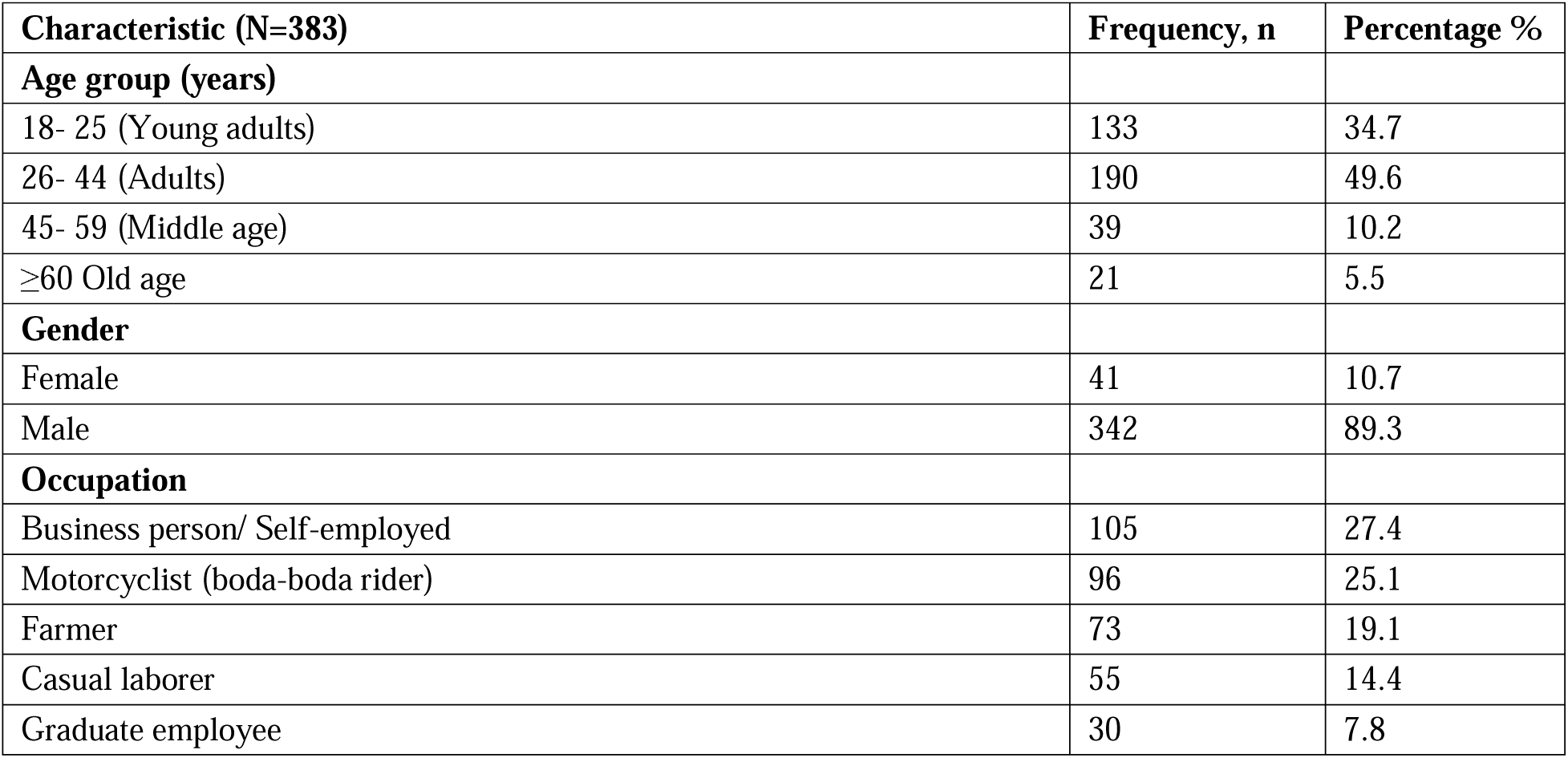
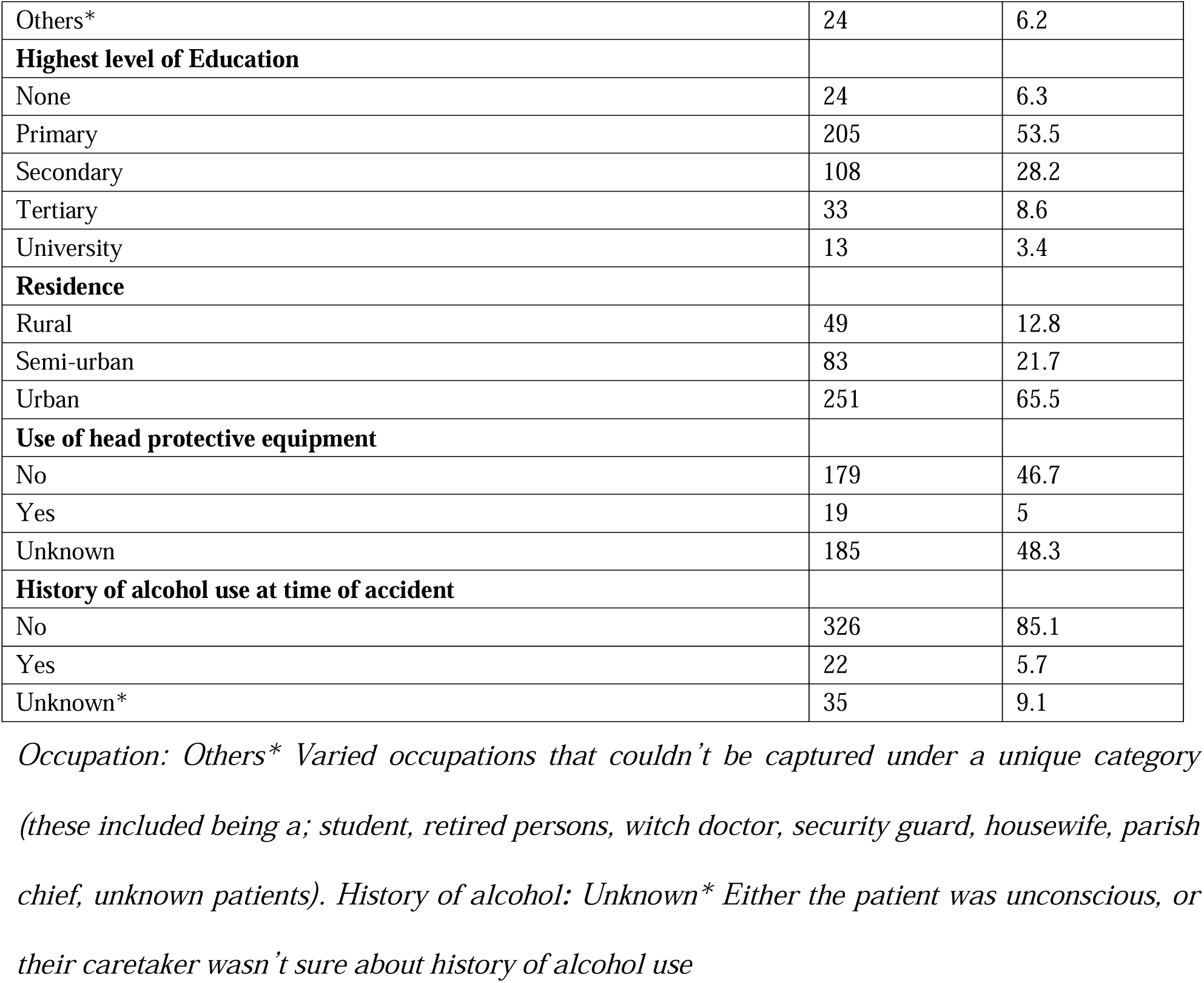
Sociodemographic characteristics of patients admitted with head injury at the Accidents and Emergency Unit of MNRH (N=383)

**Table 2:**
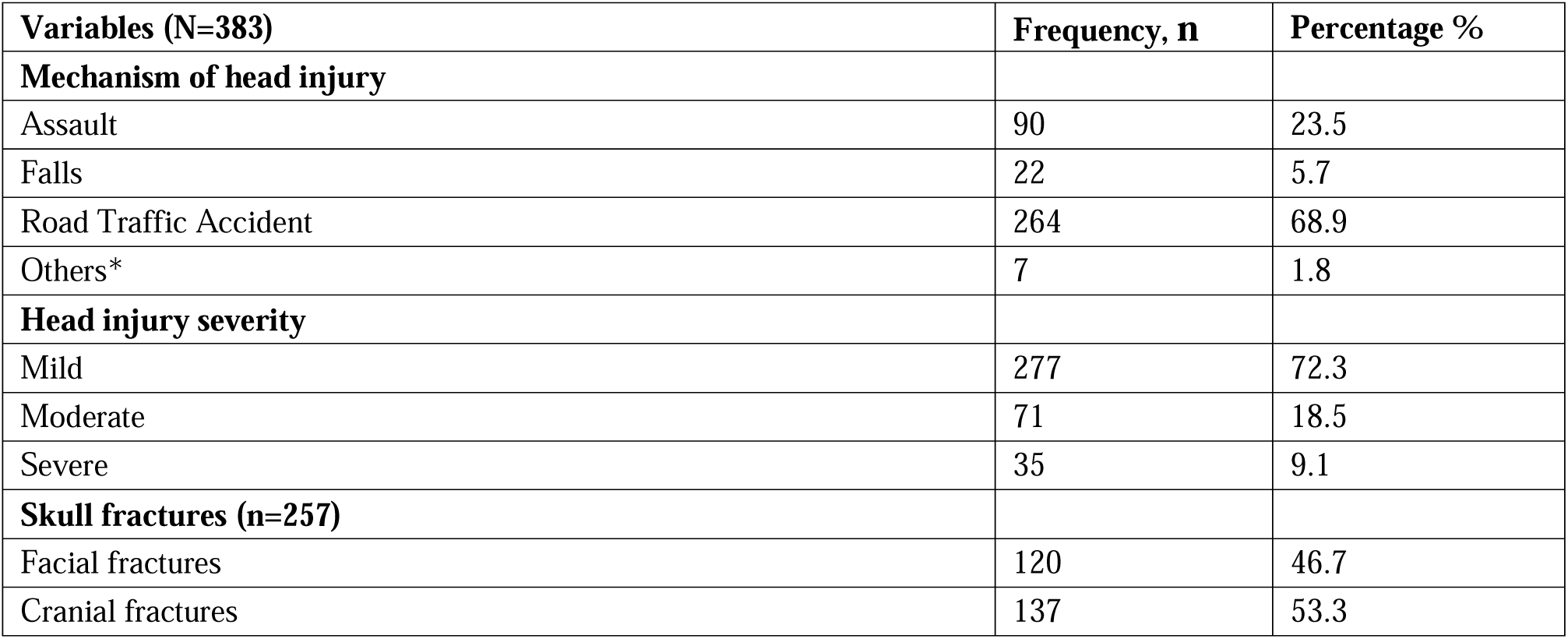
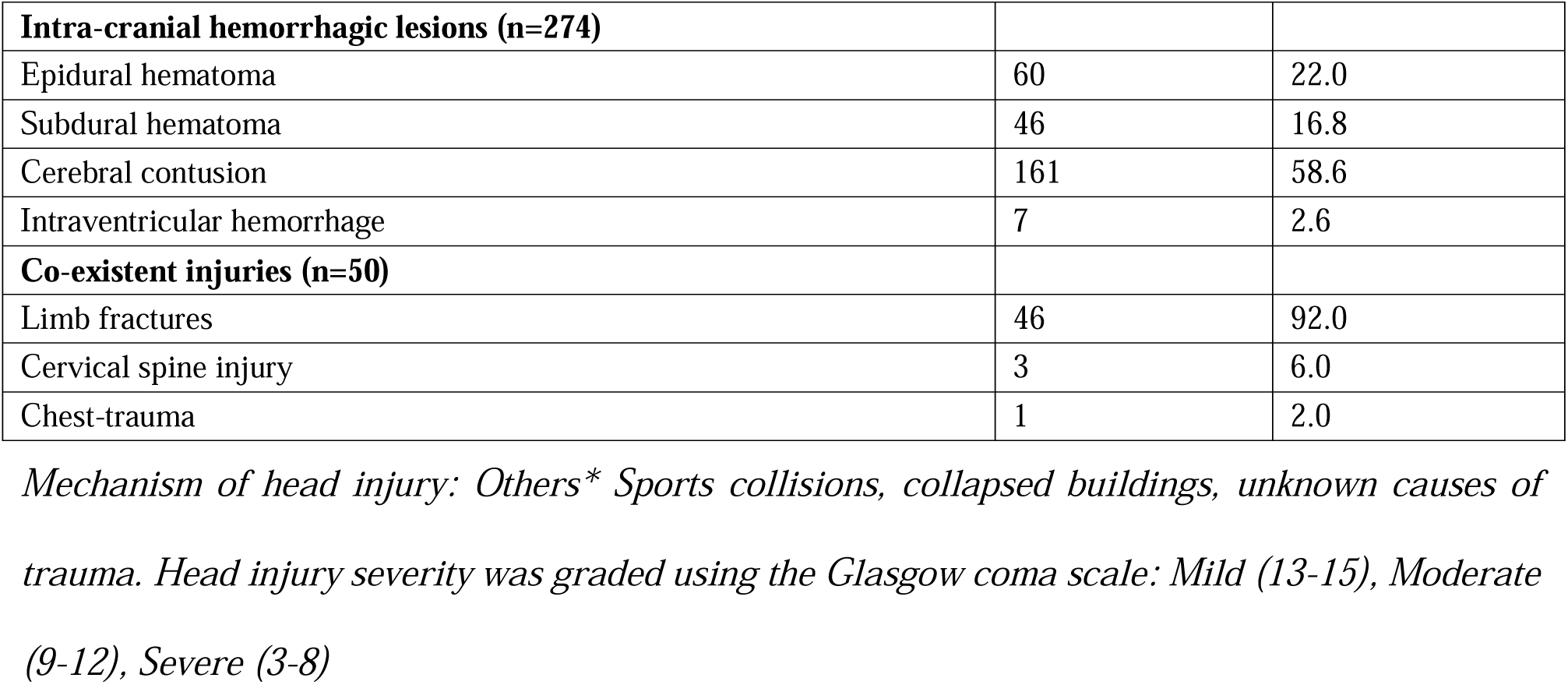
Clinical characteristics of patients admitted with head injury at the Accidents and Emergency Unit of MNRH.

### Prevalence of ocular injury

Of the 383 participants, 268 (70.0%; 95% CI 65.1-74.5) sustained at least one ocular injury (Fig 2).

**Fig 2.**
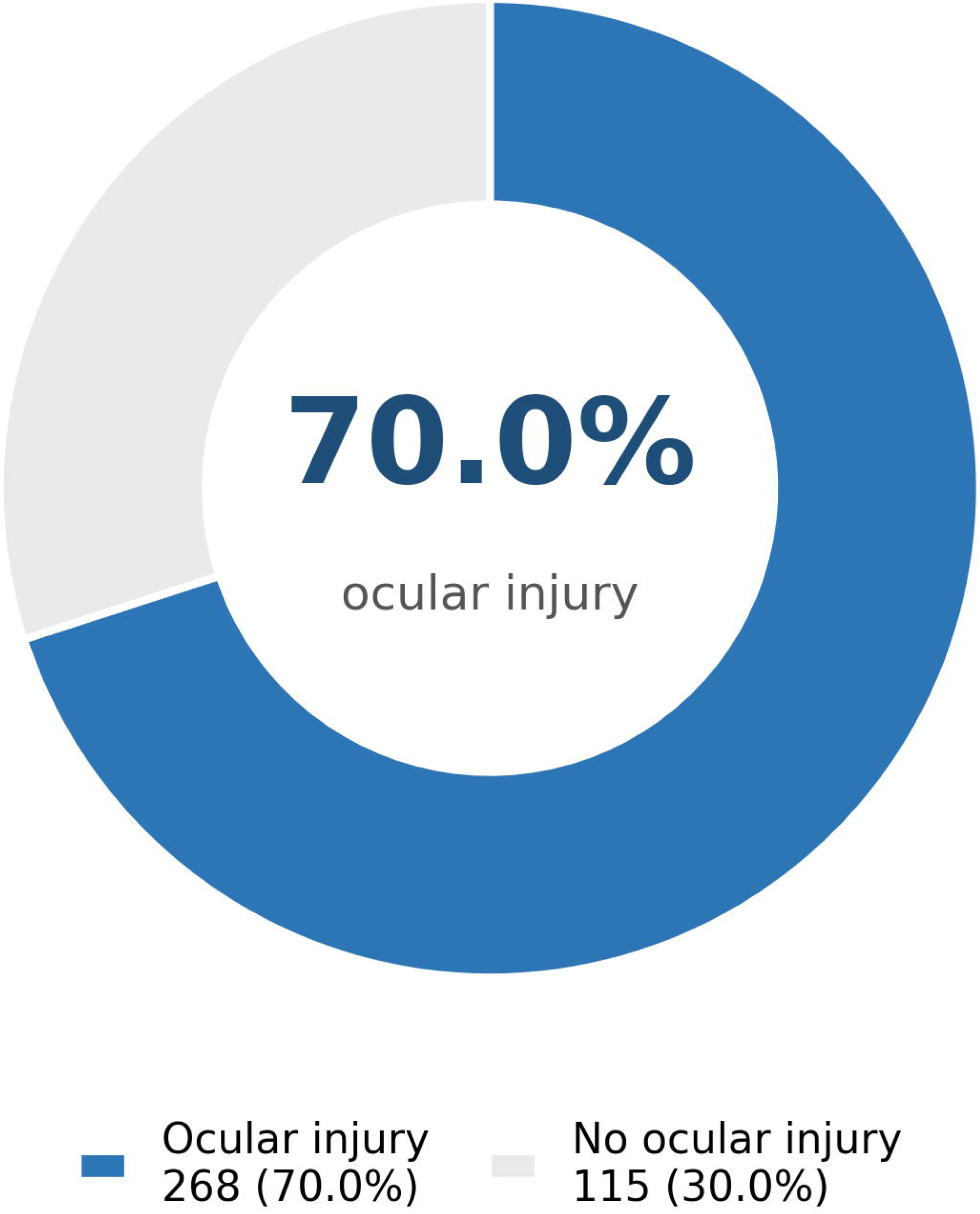
Prevalence of ocular injury among 383 patients admitted with head injury at the Accident and Emergency Unit of MNRH. At least one ocular injury was identified in 268 patients (70.0%; 95% CI 65.1-74.5).

### Patterns of ocular injury

#### Visual status at presentation

At initial bed-side assessment, normal visual acuity (≥ 6/18) was recorded in 62.1% of right eyes and 64.0% of left eyes. Approximately one third of eyes were classified as blind (< 3/60 or no light perception): 36.4% of right eyes and 36.0% of left eyes, with only small proportions showing moderate or severe visual impairment (Fig 3).

**Fig 3.**
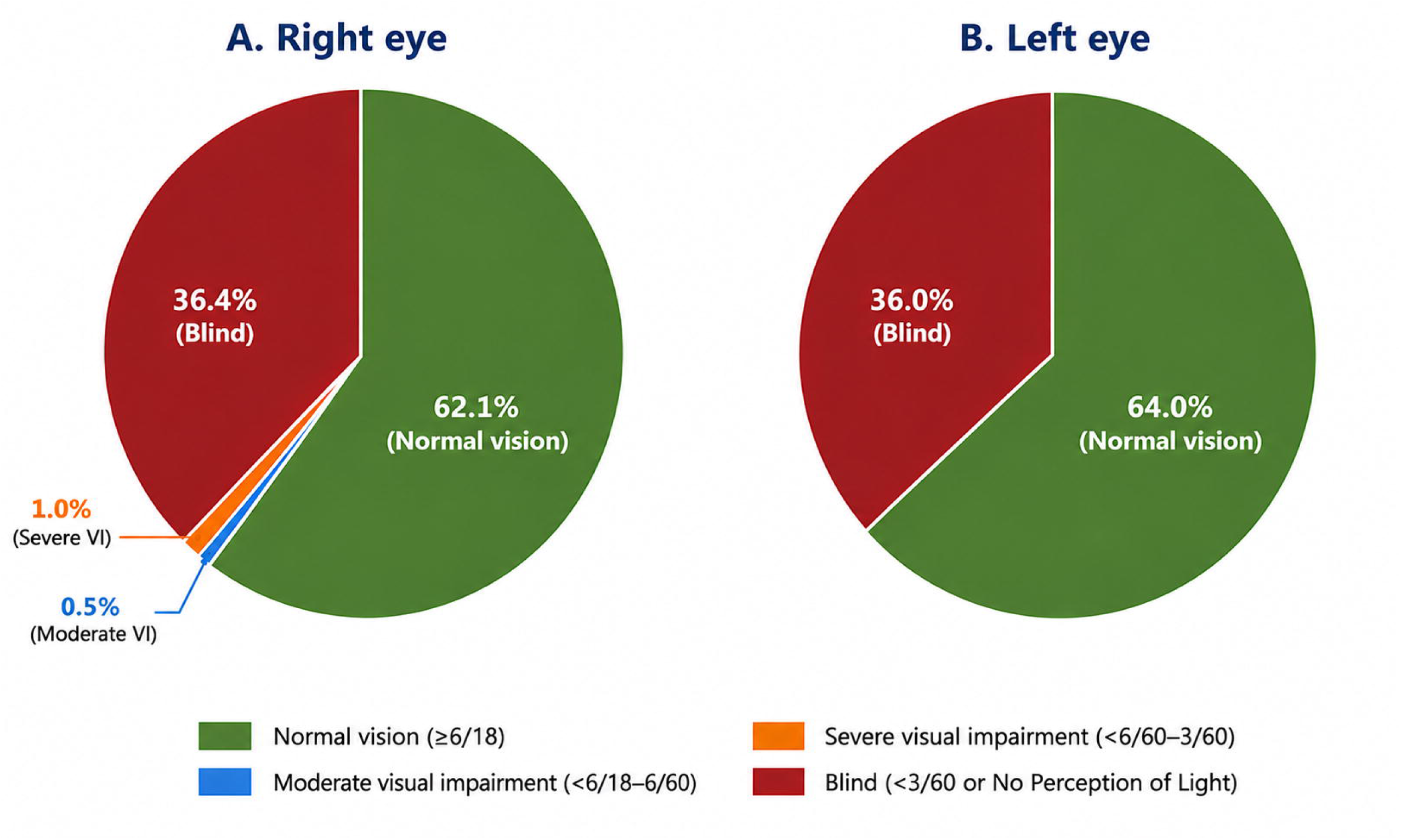
Visual status at initial presentation among patients admitted with head injury at the Accident and Emergency Unit of MNRH. Pie charts show the proportion of eyes in each World Health Organization visual-acuity category for the right eye (A) and left eye (B): normal (≥ 6/18), moderate visual impairment (< 6/18-6/60), severe visual impairment (< 6/60-3/60) and blind (< 3/60 or no light perception). In the 127 unconscious patients, visual acuity was assessed indirectly using the pupillary light reflex and recorded as light perception where a reflex response was present, or no light perception where absent.

#### Anatomical distribution

Adnexal injuries were the most prevalent category, affecting 64.0% of patients, followed by globe injuries, while neuro-ophthalmic (cranial nerve) injuries were least frequent. The most frequent individual findings are shown in Fig 4, and the full spectrum by anatomical structure is presented in Table 2.

**Fig 4.**
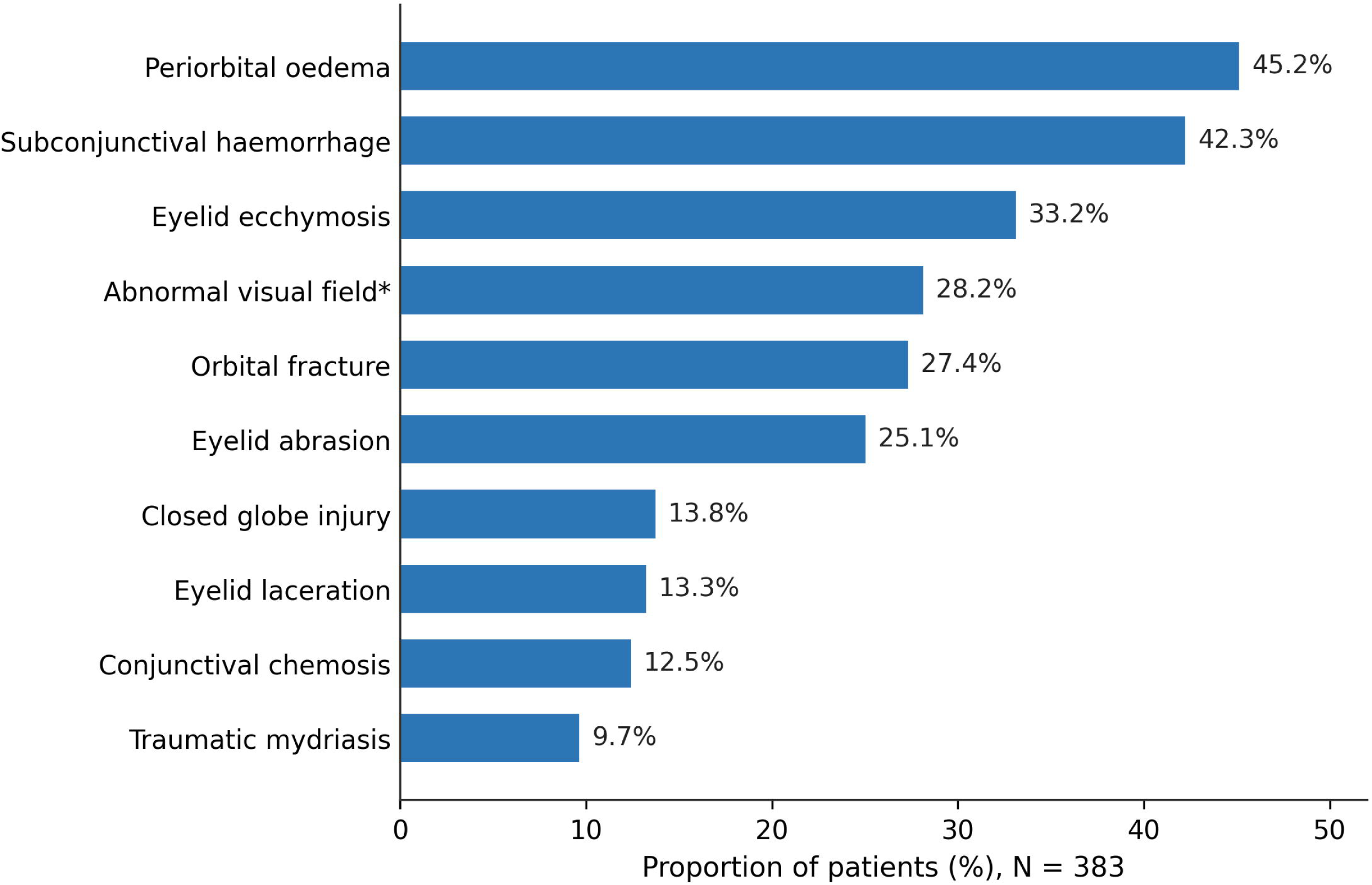
Most frequent ocular injuries among 383 patients admitted with head injury at the Accident and Emergency Unit of MNRH. Bars show the percentage of all enrolled patients with each finding. Confrontation visual fields could not be assessed in 127 unconscious patients.

#### Adnexal injuries

Adnexal injuries involved the eyelids (any eyelid injury, 57.4%), conjunctiva (43.6%) and orbit (27.9%). The most common individual findings were periorbital oedema (45.2%), subconjunctival haemorrhage (42.3%) and eyelid ecchymosis (33.2%). Orbital fractures were identified in 27.4% of patients (Table 3).

**Table 3:**
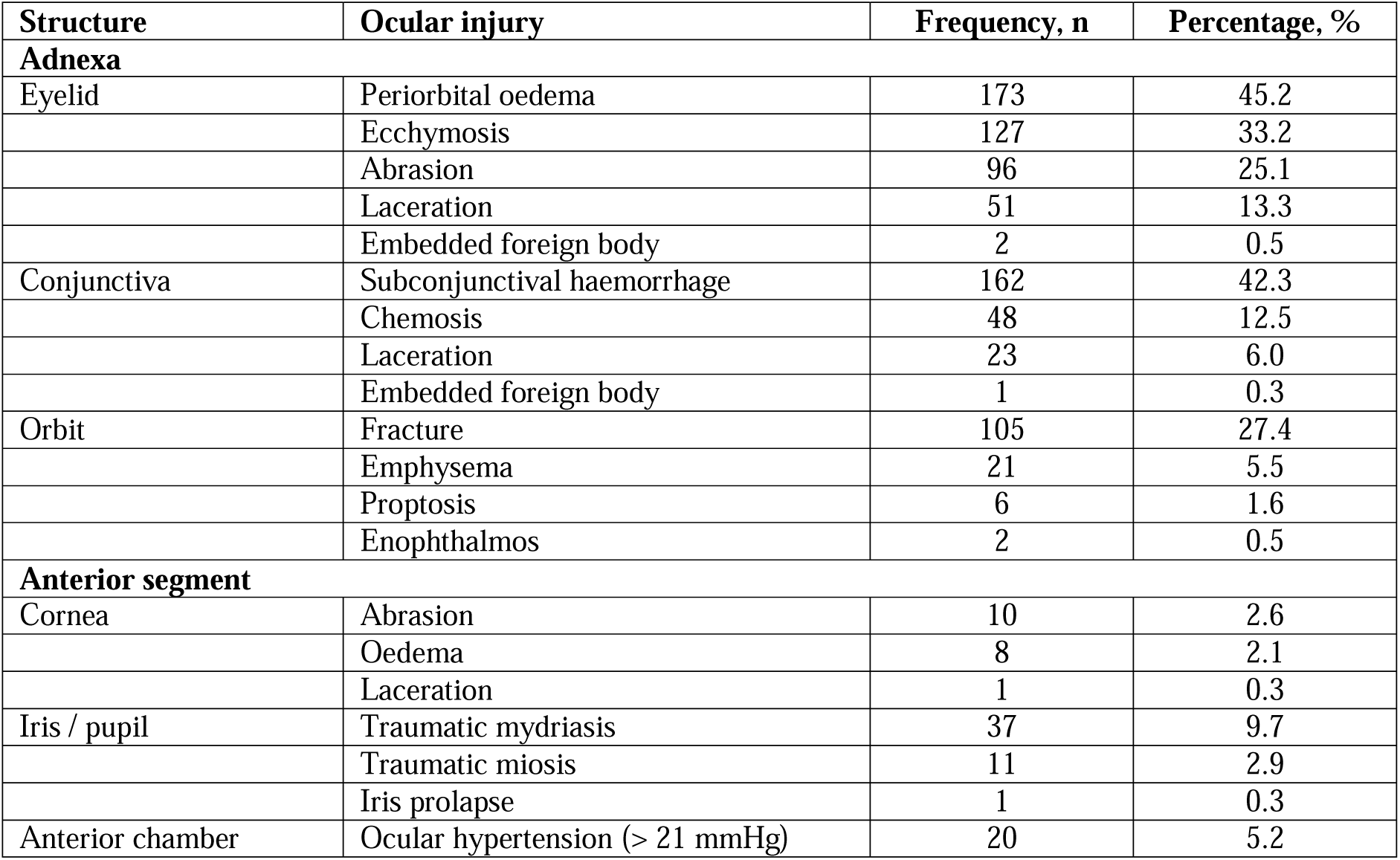
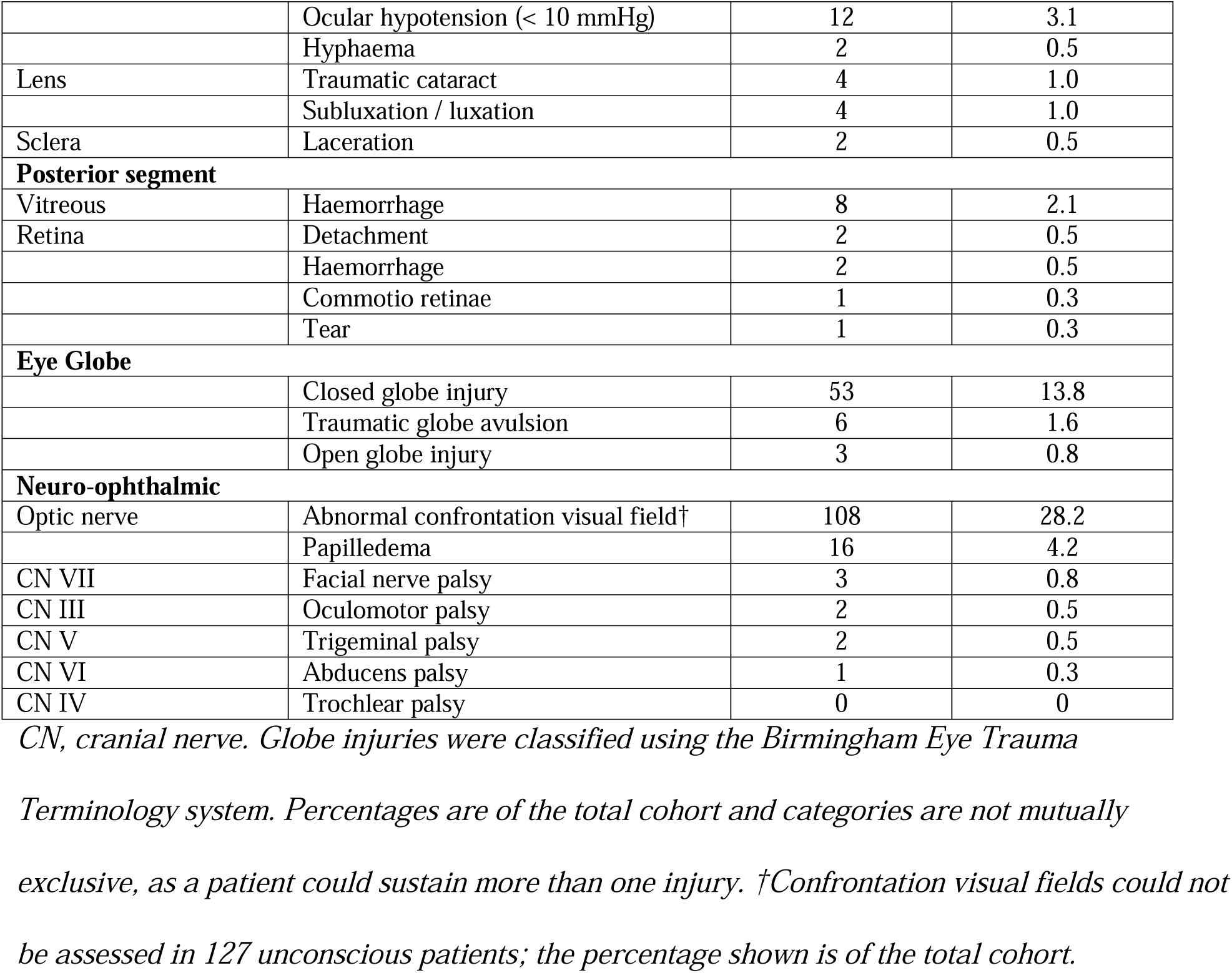
Patterns of ocular injury by anatomical structure among patients admitted with head injury at the Accidents and Emergency Unit of MNRH (N=383)

| Structure | Ocular injury | Frequency, n | Percentage, % |
| --- | --- | --- | --- |
| **Adnexa** |  |  |  |
| Eyelid | Periorbital oedema | 173 | 45.2 |
|  | Ecchymosis | 127 | 33.2 |
|  | Abrasion | 96 | 25.1 |
|  | Laceration | 51 | 13.3 |
|  | Embedded foreign body | 2 | 0.5 |
| Conjunctiva | Subconjunctival haemorrhage | 162 | 42.3 |
|  | Chemosis | 48 | 12.5 |
|  | Laceration | 23 | 6.0 |
|  | Embedded foreign body | 1 | 0.3 |
| Orbit | Fracture | 105 | 27.4 |
|  | Emphysema | 21 | 5.5 |
|  | Proptosis | 6 | 1.6 |
|  | Enophthalmos | 2 | 0.5 |
| **Anterior segment** |  |  |  |
| Cornea | Abrasion | 10 | 2.6 |
|  | Oedema | 8 | 2.1 |
|  | Laceration | 1 | 0.3 |
| Iris / pupil | Traumatic mydriasis | 37 | 9.7 |
|  | Traumatic miosis | 11 | 2.9 |
|  | Iris prolapse | 1 | 0.3 |
| Anterior chamber | Ocular hypertension (> 21 mmHg) | 20 | 5.2 |
|  | Ocular hypotension (< 10 mmHg) | 12 | 3.1 |
|  | Hyphaema | 2 | 0.5 |
| Lens | Traumatic cataract | 4 | 1.0 |
|  | Subluxation / luxation | 4 | 1.0 |
| Sclera | Laceration | 2 | 0.5 |
| **Posterior segment** |  |  |  |
| Vitreous | Haemorrhage | 8 | 2.1 |
| Retina | Detachment | 2 | 0.5 |
|  | Haemorrhage | 2 | 0.5 |
|  | Comotio retinae | 1 | 0.3 |
|  | Tear | 1 | 0.3 |
| **Eye Globe** |  |  |  |
|  | Closed globe injury | 53 | 13.8 |
|  | Traumatic globe avulsion | 6 | 1.6 |
|  | Open globe injury | 3 | 0.8 |
| **Neuro-ophthalmic** |  |  |  |
| Optic nerve | Abnormal confrontation visual field† | 108 | 28.2 |
|  | Papilledema | 16 | 4.2 |
| CN VII | Facial nerve palsy | 3 | 0.8 |
| CN III | Oculomotor palsy | 2 | 0.5 |
| CN V | Trigeminal palsy | 2 | 0.5 |
| CN VI | Abducens palsy | 1 | 0.3 |
| CN IV | Trochlear palsy | 0 | 0 |

CN, cranial nerve. Globe injuries were classified using the Birmingham Eye Trauma Terminology system. Percentages are of the total cohort and categories are not mutually exclusive, as a patient could sustain more than one injury. †Confrontation visual fields could not be assessed in 127 unconscious patients; the percentage shown is of the total cohort.

#### Globe injuries

Closed globe injuries occurred in 13.8% of patients, open globe injuries in 0.8% and traumatic globe avulsion or other destructive globe injury in 1.6%. Traumatic mydriasis was the most common anterior-segment finding (9.7%). Abnormal intraocular pressure also occurred: ocular hypertension (> 21 mmHg) in 5.2% and ocular hypotension (< 10 mmHg) in 3.1%. Posterior segment injuries were infrequent, the most common being vitreous haemorrhage (2.1%) (Table 3).

#### Neuro-ophthalmic injuries

Abnormal confrontation visual fields were detected in 108 patients (28.2% of the total cohort); testing was not possible in 127 unconscious patients. Papilledema was identified in 4.2% of patients, and in 2.1% the optic nerve head could not be visualized because of corneal or lens opacity. Cranial nerve palsies were infrequent: facial nerve (CN VII) 0.8%, oculomotor (CN III) 0.5%, trigeminal (CN V) 0.5% and abducens (CN VI) 0.3%; no trochlear (CN IV) palsy was identified (Table 3).

## Discussion

In this cross-sectional study at Uganda’s largest public tertiary referral hospital, we found a high prevalence of ocular injury (70.0%; 95% CI 65.1-74.5) among patients admitted with head injury, together with a broad spectrum of injuries spanning the adnexa, globe and neuro-ophthalmic structures. Adnexal and orbital structures were most commonly affected.

### Prevalence of ocular injury

The prevalence of ocular injury in this study (70.0%) was high and consistent with previous studies done among head injury patients in other low-resource settings. Studies from Kenya and India have also reported high prevalence rates of ocular injury ranging from 60-85% among head injury patients [18, 19]. This high prevalence is not surprising because the possibility of having ocular injury among patients with head injury is increased by the proximity of the visual system in the head. Traumatic forces are directly transmitted from the skull bones via the orbital bones to the eye structures, hence increasing the possibility of ocular injury [6]. Earlier studies at MNRH reported lower prevalences of 35-52.5% [13, 14], but these were restricted to specific injury mechanisms or did not include comprehensive ophthalmic assessment with tonometry and posterior-segment evaluation. The higher prevalence in our study probably reflects the high-risk patient population (head injury), inclusive enrolment of all head injury mechanisms, combined with a thorough, comprehensive, examination protocol. The much lower prevalence rates of 1-2.2% reported from high-income countries [20, 21] likely reflect differences in road safety enforcement as well as the availability and routine use of protective gear in high income settings; in our cohort, only 5% of patients were documented to have used head protection and road traffic accidents, which involve high energy transfer, accounted for 68.9% of cases.

### Patterns of ocular injury

#### Visual status

Although most patients retained normal measured acuity in at least one eye, approximately one third of eyes were classified as blind at bed-side assessment on presentation, a higher proportion than the 29.2% reported by *Masila et al* in Kenya [18]. This difference is most plausibly explained by the timing of assessment: we measured visual acuity at first contact during the acute trauma phase, when periorbital oedema, ocular-surface injury, pain and reduced consciousness can all confound measurement. Acute bedside acuity testing is known to underestimate eventual visual function [22], so these figures should be interpreted as indicators of early functional status rather than definitive visual outcomes.

#### Adnexal injuries

Adnexal injuries predominated, consistent with studies from Uganda, Kenya and Nigeria that identify adnexal structures as the most frequently injured ocular structures following head trauma [13, 18, 23]. This is anatomically predictable, as the periorbital soft tissues and orbital rim act as the primary buffer for external forces during craniofacial trauma [24].

#### Globe injuries

Closed globe injuries were more common than open globe injuries (13.8% versus 0.8%), in keeping with the worldwide literature: blunt trauma mechanisms predominantly produce closed globe injuries, whereas open globe injuries (often due to penetrating trauma, or industrial accidents) are less frequent but carry a poorer visual prognosis [15, 16]. Series across Sub-Saharan Africa similarly report closed globe injuries as the majority of presentations [25, 26]. Traumatic globe avulsion (1.6%) is a rare but devastating injury consistent with the severity of the road traffic collisions and cases of assault seen in this setting.

#### Anterior and posterior segment injuries

14.88% of the patients had at least one anterior segment injury. These injuries included traumatic mydriasis (9.7%), traumatic miosis (2.9%), corneal abrasion (2.6%), lens luxation (1%), hyphema (0.5%) and corneal laceration (0.3%). Some of these patterns are consistent with those reported by *Devandan et al* [27]. Hyphema in particular, though occurring infrequently, is clinically important because it is a marker of significant globe contusion and predicts raised intraocular pressure and risk for secondary glaucoma.

The largely normal intraocular pressure measurements seen may be partly attributable to the routine use of intravenous mannitol or hypertonic saline for osmotherapy in head injury patients, which also lowers intraocular pressure [28]. Among patients noted with intraocular pressure changes (raised intraocular pressure in 5.2% and hypotony 3.1% in our cohort), other studies have recognized these changes as early indicators of anterior-segment trauma [29, 30].

Posterior segment injuries were found in 16.45% of the patients in this study. These injuries included vitreous hemorrhage in 2.1%, commotio retinae in 0.3% and retinal detachment in 0.5% of cases. These findings were similar to those in a study by *Devandan et al* among RTA victims, although among a smaller cohort of 67 participants [27]. *Devandan et al* described vitreous hemorrhage in 9% of the participants, retinal detachment in 1.5% and commotio retinae in 9%. *Wold et al* found a higher prevalence of 50.8% of posterior segment injuries. However, the mechanism of injury in their group was commonly an improvised explosive device [31].

#### Neuro-ophthalmic injuries

Cranial nerve palsies (III, V, VI, VII) were observed infrequently but were clinically significant when present. 3 patients with facial nerve palsy presented with lagophthalmos, while the 2 with Trigeminal nerve palsy noted at assessment exhibited reduced periocular skin sensation and reduced corneal sensation. These palsies are more commonly associated with skull base or orbit apex fractures and high energy mechanisms. Traumatic optic neuropathy (TON) was uncommon at presentation but remains an important delayed complication that requires follow-up. This mirrors an observation by *Blanch et al* that while TON occurs in 0.5-8% of civilian traumatic brain injury cases, subclinical or undiagnosed optic nerve damage is much more common [32]. Abnormal confrontation visual fields were detected in 28.2% of the cohort, but this must be interpreted cautiously: testing was impossible in 127 unconscious patients, and in the acute setting of trauma, periorbital swelling and poor cooperation may produce apparent deficits that do not reflect true neurological loss [33]. Repeat perimetry after swelling resolution is essential to differentiate true neurological loss from mechanical obstruction. Papilledema (4.2%) is consistent with raised intracranial pressure complicating severe head injury[34].

### Strengths and limitations of this study

Strengths of this study include a relatively large sample size, the inclusion of both conscious and unconscious patients to minimize selection bias, and a comprehensive standardized examination protocol-with tonometry, posterior-segment and neuro-ophthalmic assessment, and radiological correlation, which were lacking in prior Ugandan studies.

Several limitations should be acknowledged. As a single-centre study at a tertiary referral hospital that receives predominantly severe trauma cases, the observed prevalence may overestimate that in lower-level or community settings. Ophthalmic assessment was performed only at admission, precluding evaluation of visual prognosis or the evolution of injury. Acute visual acuity testing and confrontation visual field assessment could be limited by altered levels of consciousness among some patients, poor cooperation and periorbital swelling in the acute setting of trauma, potentially inflating the proportion classified as visually impaired. Visual acuity in unconscious patients was estimated using the pupillary light reflex as a proxy for light perception; because this reflects a brainstem pathway rather than confirmed cortical visual perception, it may misclassify visual status in patients with isolated cortical or optic-pathway injury. Self-reported data on helmet use and alcohol consumption are subject to recall and social-desirability bias, and the cross-sectional design limits causal inference.

## Conclusions

Ocular injury is highly prevalent among patients admitted with head injury at a tertiary referral hospital in Uganda, affecting seven in ten patients, with adnexal and orbital structures most commonly involved, followed by globe and neuro-ophthalmic injuries. These findings support the integration of routine ophthalmic screening into the initial evaluation of all head injury patients in this setting, alongside timely referral for ophthalmic care.

## Supporting information

Data set

## Data Availability

The minimal data set has been attached as a supporting file of this manuscript.

## Acknowledgments

The authors thank the staff of the Accident and Emergency Unit and the Department of Ophthalmology at Mulago National Referral Hospital, and the research assistants, for their support during data collection. We are grateful to the study participants and their families.

## Supporting information

**S1 Data set for patients admitted with head injury at MNRH**

